# Distinguishing syndromic and nonsyndromic cleft palate through analysis of protein-altering de novo variants in 816 trios

**DOI:** 10.1101/2025.03.03.25323233

**Authors:** Kelsey R. Robinson, Sarah W. Curtis, Justin E. Paschall, Terri H. Beaty, Azeez Butali, Carmen J. Buxó, David J. Cutler, Michael P. Epstein, Jacqueline T. Hecht, Lina Moreno Uribe, Gary M. Shaw, Jeffrey C. Murray, Harrison Brand, Seth M. Weinberg, Mary L. Marazita, Kimberly F. Doheny, Elizabeth J. Leslie-Clarkson

## Abstract

*De novo* variants (DNs) are sporadically occurring variants that most commonly arise in the germline and are present in offspring but absent in both parents. As they are not under selective pressure, they may be enriched for disease-causing alleles and have been implicated in multiple rare genetic disorders. Cleft palate (CP) is a common craniofacial congenital anomaly occurring in ∼1 in 1700 live births. Genome-wide association studies for CP have found fewer than a dozen loci, while exome and targeted sequencing studies in family-based and case-control cohorts often lack statistical power to conclusively identify causal genes. Based on previous work by our group and others, deciphering the genetic architecture of CP and gene discovery efforts are complicated by the heterogeneous nature of the disorder. We aggregated sequence data for 816 case-parent trios with CP, representing all subtypes of CP and roughly evenly split between isolated and syndromic presentations. We hypothesized there would be a burden of DNs in CP probands and tested this hypothesis in the full cohort and various phenotypic subgroupings. We identified global enrichment of protein-altering DNs (1.36, p=2.39×10^-22^), and exome-wide significant (p<1.3×10^-6^) gene-specific enrichment for *SATB2*, *MEIS2*, *COL2A1*, *ZC4H2*, *EFTUD2*, *KAT6B*, and *ANKRD11.* We found a statistically significant higher enrichment of loss-of-function and missense DNs in syndromic (1.49, p=2.84×10^-19^) versus nonsyndromic probands (1.25, p=4.01×10^-7^) but no differences between CP subtypes. We also evaluated biological differences, identifying distinct enrichments across two single cell RNA sequencing datasets: mouse palate at the time of palate fusion and human embryos at post-conceptional weeks 3-5. Altogether, we show DNs are a contributor to CP risk, and that combined analysis can enhance our ability to find genetic associations that would otherwise be undetected.

## Introduction

Cleft palate (CP) is one of three major subtypes of orofacial clefts (OFCs). Together with those affecting the upper lip (cleft lip with or without cleft palate, CL/P), OFCs are the most common craniofacial birth defect. CP occurs in approximately 1 in 1700 live births, though there are differences in geographical and ancestral frequencies (1). In the United States, approximately 2,600 babies are born with overt CP each year and up to 80,000 babies are born with the more subtle submucous cleft palate (SMCP) (2). The secondary palate, or the roof of the mouth, separates the oral and nasal cavities and is made up of two primary components: an anterior bony hard palate that facilitates normal feeding, and a posterior muscular soft palate that elevates to close off the pharynx during swallowing and speech. Left untreated, CP is often fatal early in life due to aspiration and/or malnutrition from feeding difficulties (3) though prognosis is much more favorable with surgical correction. However, individuals born with CP often go on to face speech and/or hearing problems, require advanced orthodontic care, and can experience additional comorbidities as they age (4, 5). As such, CP creates both individual and public health burdens. Improving our understanding of its origin can lead to enhanced prevention, treatment, and prognosis for affected individuals.

Despite approximately 25% of CP affected individuals having a positive family history (6, 7), identification of genetic variants causing or increasing risk for CP remains elusive. Collectively, fewer than a dozen associated loci have been discovered with genome-wide association studies (GWAS) for CP (8–14). Genes such as *IRF6* (8), *GRHL3* (9)*, and CTNNA2* (10) have been implicated, though these associations are often population-specific. One study in a Chinese population identified 9 associated loci (12) but many were not replicated in a subsequent GWAS in an independent Chinese population (11). Given a relative lack of common variant associations with CP, another avenue of discovery is the contribution of rare variants.

A rare variant hypothesis is further supported by the fact that CP occurs as part of a syndrome in about 50% of occurrences, compared to 30% in CL/P. Further, we previously found individuals with CP were more likely to have rare pathogenic or likely pathogenic (P/LP) variants in genes assembled from clinical genetic testing panels than other types of OFCs (15). Specifically, we found the diagnostic yield for 58 CP trios was 18% compared to 9% in CLP and 3% for CL. This suggests that CP may more frequently have a monogenic cause than CL/P. In comparison, a more comprehensive evaluation of rare variants in 603 syndromic OFC probands by Wilson et al. (16) found a diagnostic yield for P/LP variants was 36.5%, though the difference between subtypes was less dramatic: 33.3% for CL/P and 37.1% for CP. Among the P/LP variants with known inheritance information, 73% were *de novo,* indicating these types of variants are great candidates for additional investigation.

Our previous work has shown nominal significance for coding *de novo* variants (DNs) in 58 CP trios (enrichment 1.39, p=9.32×10^-3^), but lacked power to identify new genes associated with CP (17), indicating a need for larger studies. In addition, we have shown that phenotypic heterogeneity can mask novel genes and hamper gene discovery (14). Therefore, we assembled a phenotypically and ancestrally diverse cohort of 816 CP trios from the CPSeq, Gabriella Miller Kids First, and Deciphering Developmental Disease (DDD) studies. We also explored the heterogeneous nature of CP by investigating our cohort stratified by proband sex, CP subtype (involvement of the hard versus soft palate), and by the presence or absence of additional non-cleft phenotypic features.

## Methods

### CPSeq Dataset

We assembled a collection of 473 case-parent trios ascertained on proband affection status (e.g., cleft palate) from the CPSeq (N=429) and Gabriella Miller Kids First (GMKF) (N=44) whole genome sequencing projects (17) (hereafter referred to as CPSeq). There were 14 samples that underwent WGS in both studies, so duplicates were removed prior to analysis to ensure an individual was only included once. Trios represent all major ancestry groups affected by CP including those with European ancestry (recruited from Spain, Turkey, Hungary, United States), Latin America (Puerto Rico, Argentina), Asia (China, Singapore, Taiwan, the Philippines), and Africa (Nigeria, Ghana). Recruitment and phenotypic assessment occurred at multiple domestic and international sites following institutional review board (IRB) approval for each local recruitment site and coordinating centers (University of Iowa, University of Pittsburgh, and Emory University).

There were 278 female probands and 195 male probands. All probands and parents were assessed for the presence of a CP with ∼2/3 of the assembled samples undergoing additional phenotyping to assess the location and severity of the CP. There were 140 probands with cleft hard and soft palate (CH&SP), 164 cleft soft palate (CSP), 5 cleft hard palate (CHP), 27 submucous cleft palate (SMCP), and 137 with unspecific CP subtypes. Although probands/trios were not excluded based on additional clinical features consistent with a syndromic diagnosis, only 33 trios were classified as possibly or probably syndromic based on a reported presence of additional major or minor clinical features. Additional breakdown based on genetic ancestry and subcategories is available in **Supplemental Table 1**.

### Deciphering Developmental Disease (DDD) Dataset

We accessed the publicly available DDD data which is primarily ascertained based undiagnosed neurodevelopmental disorders (NDDs) and/or congenital anomalies, abnormal growth parameters, dysmorphic features, and unusual behavioral phenotypes (18) using the 2017-15-12 data freeze. We used the published list of DNs detailed in Kaplanis, et al. (19) which includes data from 9,858 trios. Using a list of HPO terms related to clefting (**Supplemental Table 2**), we identified 346 probands with cleft palate and/or bifid uvula. There were 193 males and 153 females with CP. The DDD cohort was ascertained from regional genetics services in the UK and Ireland.

### Whole genome sequencing

The full description of sequencing and variant calling methodology for the CPSeq trios is detailed in Robinson et al. (14) and for the GMKF trios in Bishop et al (17). The WGS for CPSeq was performed by the Center for Inherited Disease Research (CIDR) at Johns Hopkins University (Baltimore, MD). The DRAGEN Germline v3.7.5 pipeline on the Illumina BaseSpace Sequence Hub platform was used for alignment, variant calling, and quality control, resulting in a single multisample VCF file. For GMKF, WGS for European samples was carried out by the McDonnell Genome Institute (MGI) the Washington University School of Medicine (St. Louis, MO) followed by realignment to hg38 and variant calling at the GMKF Data Resource Center at the Children’s Hospital of Philadelphia. WGS for Colombian and Taiwanese samples was carried out by the Broad Institute, with alignment to hg38 and variant calling by GATK pipelines (20–22).

### Identification of *de novo* variants

The DRAGEN 3.7.5 aligner and variant caller was used to generate gVCF files for each CPSeq trio. Individual trio VCFs with *de novo* variant tags were then generated by using gVCF files combined with pedigree information as input to the DRAGEN 3.7.5 joint caller. To be considered a DN, variants had to have a quality score of 30 and DQ>2.The pipeline for DN called in the GMKF cohort is detailed in Bishop et.al (17). For both cohorts, genotypes were set to missing if GQ<20 or read depth <10, and parental genotypes had to be confirmed homozygous reference (0/0), pass all filtering steps, and have an allele balance (AB) ratio of <0.05.

### Variant Annotation

Variants were annotated with ANNOVAR (version 201910). Variant with coding consequences were selected based on classification as “exonic” or “splicing”, and only variants with MAF of <0.5% in either gnomAD exomes v2.1.1 or gnomAD v3.1.2 were retained for analysis.

### DN enrichment

We evaluated coding DN enrichment using the R package ‘DenovolyzeR’ package (version 0.2.0). After comparison of genes with DNs and genes with mutation rates (obtained using the function ‘viewProbabilityTable()’), we analyzed 471 case-parent trios as there were 2 trios with DNs in genes not present in the table. The cohort was tested for an excess of DNs exome-wide and per gene using the functions ‘DenovolyzeByClass’ and ‘DenovolyzeByGene’, respectively. These functions utilize mutation models described by Samocha, et al. (23) to determine if there are more observed DNs in a dataset than would be expected by chance. Using mutational rates, the number of DNs is expected to follow a Poisson distribution under the null model of no association between a variants class and a phenotype

(24). Given a fixed sample size and expected mutation rate for a given genetic sequence, we can determine M (both the mean and variance) as well as the standard deviation, resulting in the “known” constant. Under the alternate model, the number of observed mutations, A, also follows a Poisson distribution, but A may not equal M. We used the Poisson distribution to determine enrichment shown by A/M with 95% confidence intervals.

For the DenovolyzeByGene analysis, we used a multiple test correction for the number of genes with DNs and considered genes significant at p<5 ×10^-5^ (0.05/1000 genes with DNs). We also considered a more conservative threshold for exome-wide significance at p<1.30×10^-6^. In analyses where we were interested in the enrichment in specific genes sets, the function ‘includeGenes’ was applied, and we corrected for the number of clusters tested. We also tested for significant differences between DN enrichment in males and females using a Z test for the observed versus expected number of variants in males versus females while considering the prevalence differences between sexes. We assumed the variance of observed variants was equal to the expected variance, based on the Poisson distribution, and determined Z using (observed – expected) / sqrt (expected) for each class of variant.

### Enrichment analyses and creation of gene sets

We evaluated our dataset for enrichment in several different ways. First, we performed a gene set enrichment analysis (GSEA) on all protein-altering DNs using the freely available webserver gProfiler

(25). Next, we created three sets of genes directly relevant to CP: an OFC-specific gene panel (15), a set of marker genes generated from single nucleotide RNA sequencing (snRNAseq) of the secondary palate in mice at embryonic day 15.5 (E15.5) (26), and a set of marker genes from human embryos at post conception weeks 3-5 (27). The OFC gene panel was curated from four sources, including the National Health Service (NHS) Genomic Medicine Service cleft panel (v2.2), the Prevention Genetics CL/P clinical genetic testing panel, genes from the Online Mendelian Inheritance in Man (OMIM) that include OFCs with a known inheritance and molecular basis, and a manually curated list from recent research studies on OFC genetics—additional details on curation are published in Diaz Perez, et al (15). The full details on the mouse snRNAseq and marker gene generation can be found in Piña, et al.

(26). We filtered marker genes for FDR <0.01 prior to enrichment testing for DNs. The full details for marker gene generation for the human embryos snRNAseq can be found in Zeng, et al. (27).

## Results

### Cleft palate probands are enriched for *de novo* variants

We evaluated exome-wide enrichment of *de novo* variants (DNs) from a starting dataset of 819 cleft palate case-parent trios. These were made up of 473 probands ascertained on CP and 346 probands ascertained primarily based on undiagnosed developmental disorders as part of the Deciphering Developmental Disorders (DDD) study (16).

We identified 1,121 protein-coding variants in 1,015 genes (**Supplemental Table 3**), averaging 1.37 DNs per trio (**Supplemental Figure 1A**). DN frequency followed a Poisson distribution with no significant deviation tested by chi-square goodness-of-fit (p=0.83). There was, however, a higher rate of DNs among syndromic probands, with 79% of syndromic versus 71% of non-syndromic probands having at least 1 coding DN. This resulted in a statistically significant increase in the DN rate averaging 1.49 and 1.24 per syndromic and nonsyndromic trio, respectively (p=0.005).

We classified DNs based on the variant type and predicted function as follows: synonymous variants, missense variants (including single amino acid substitutions and in-frame insertions or deletions), putative loss-of-function variants (pLOF, including nonsense, frameshift insertions or deletions, and splice acceptor or donor sites), and a category referred to as protein-altering (PA) variants, which includes the combination of all missense and pLOF variants. Broken down into these categories, we had 233 synonymous, 721 missense, 167 pLOF, and a combined 888 PA DNs (**Supplemental Figure 1B**). Because we utilized the built-in mutation rates for denovolyzeR, there were 15 genes with DNs but without expected mutation rates (6 synonymous, 9 missense). In cases where probands only had DNs in these genes, they were removed from analysis, resulting in 816 effective trios with 1106 DNs (167 synonymous, 712 missense, 227 pLOF) undergoing full evaluation.

We first tested DN enrichment in all CP trios using denovolyzeR on an exome-wide basis. CP probands had significantly more coding DNs (1.36, p=2.39×10^-22^) than would be expected by chance based on mutational models (23). When split by variant class, there was no enrichment of synonymous variants (0.99, p=0.56), which is expected for two reasons: synonymous variants are not often causal for disease, and the lack of enrichment indicates there is no overall increased rate of variation in our samples. There was, however, significant enrichment of PA variants (1.50, p=1.24×10^-29^), driven by both missense (1.38, p=1.24×10^-16^) and pLOF (2.34, p=3.91×10^-22^) variant classes (**Figure 1A**, **Supplemental Table 4**). When comparing variant classes, the main difference between syndromic and nonsyndromic probands was the number of pLOF DNs with enrichments of 3.15 (p=6.22×10^-23^) versus 1.66 (p=1.23×10^-4^), respectively (**Figure 1B**). The difference in pLOF variants was statistically significant (p=6.33×10^-5^), but there were no significant differences for synonymous or missense variants.

**Figure 1:**
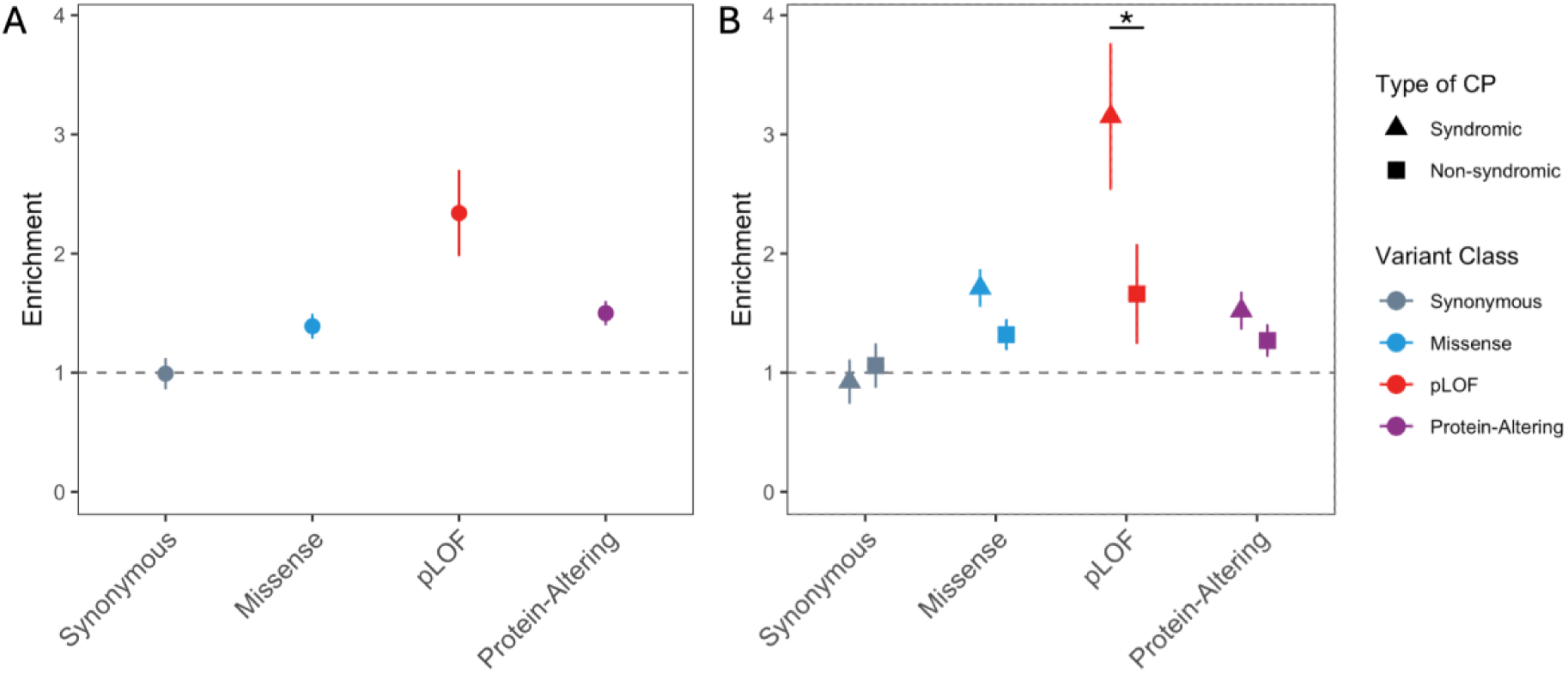
CP proband are enriched for protein-altering *de novo* variants with denovolyzeR. A) Exome-wide enrichment for DNs in 816 CP probands. B) Comparison of enrichment for syndromic (N=378) and nonsyndromic (N=434) probands. The horizontal dotted line at 1 represents no enrichment (where observed = expected). *p=6.33×10^-5^

We next compared male versus female probands, as females are more frequently affected by CP. The combined CP cohort had a female bias (M:F ratio of 0.9), which was more pronounced in nonsyndromic probands (M:F ratio 0.7). Interestingly, syndromic probands were male-biased (M:F ratio 1.12). We compared our data to ratios reported from EUROCAT (28), and found our ratios for the full cohort or nonsyndromic probands was consistent with their registry-based ratios (p>0.05); however, we found the M:F ratio of our syndromic probands significantly differed from their reported ratio of 0.89 (p=0.017). When evaluating differences by DN enrichment within the cohort, we found both sexes were enriched for PA variants (male 1.59, p=6.80×10^-20^; female 1.42, p=2.29×10^-12^), and there were no significant differences between sex for the full cohort or by syndromic status (**Figure 2**, **Supplemental Table 5**). Therefore, DNs do not appear to explain the sex bias typically observed in CP.

**Figure 2:**
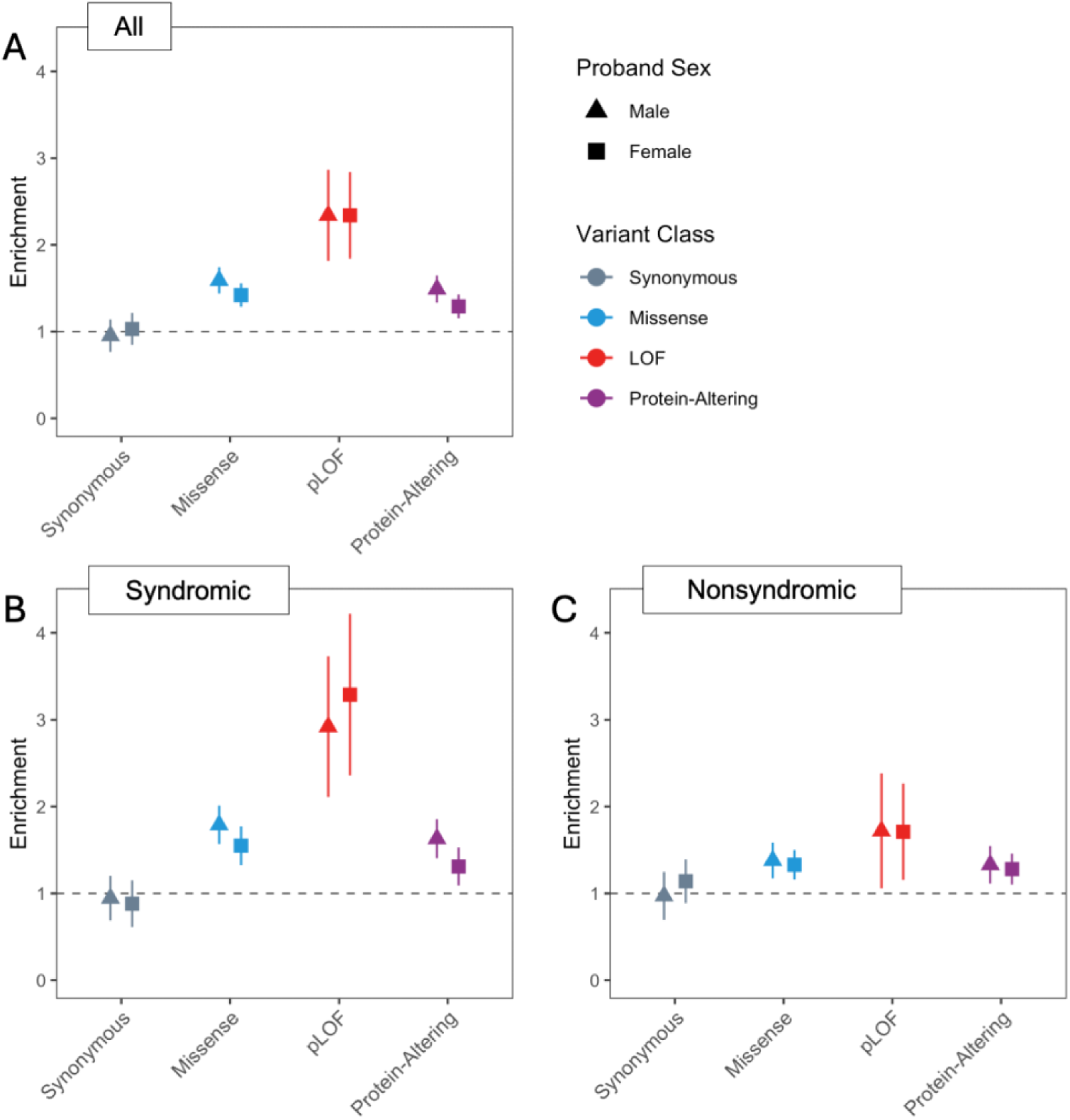
No differences in DN enrichment is observed between proband sex. A) Exome-wide enrichment for DNs in 386 male and 430 female CP probands. B) Enrichment for 204 male and 174 female syndromic CP probands. C) Enrichment for 180 male and 254 female nonsyndromic CP probands. The horizontal dotted line at 1 represents no enrichment (where observed = expected).

We compared enrichment by CP subtype for cleft hard and soft palate (CH&SP, n=139), cleft soft palate (CSP, n=199), and submucous cleft palate (SMCP, n=86), based on the rationale that DNs may be more prevalent in more “severe” forms of CP (**Figure 3**, **Supplemental Table 4**). Overall, the PA enrichment for each subtype was similar (CH&SP 1.45, p=1.24×10^-5^; CSP 1.45, p=2.69×10^-7^; SMCP 1.68, p=3.55×10^-5^) and there were no differences in variant class enrichment. This was not the case for pLOF variants, which were significantly higher in SMCP compared to both CSP (p=0.043) and CH&SP (p=0.003). However, this is most likely explained by an ascertainment bias as isolated SMCP is more likely to be identified in patients undergoing a full clinical workup and ascertained for other conditions, rather than identified as an isolated phenotype. Accordingly, the majority (70%) of our SMCP probands were in the syndromic group, which we know also is significantly more enriched for pLOF DNs. Altogether, there does not seem to be a relationship between DN enrichments or DN class and CP severity.

**Figure 3:**
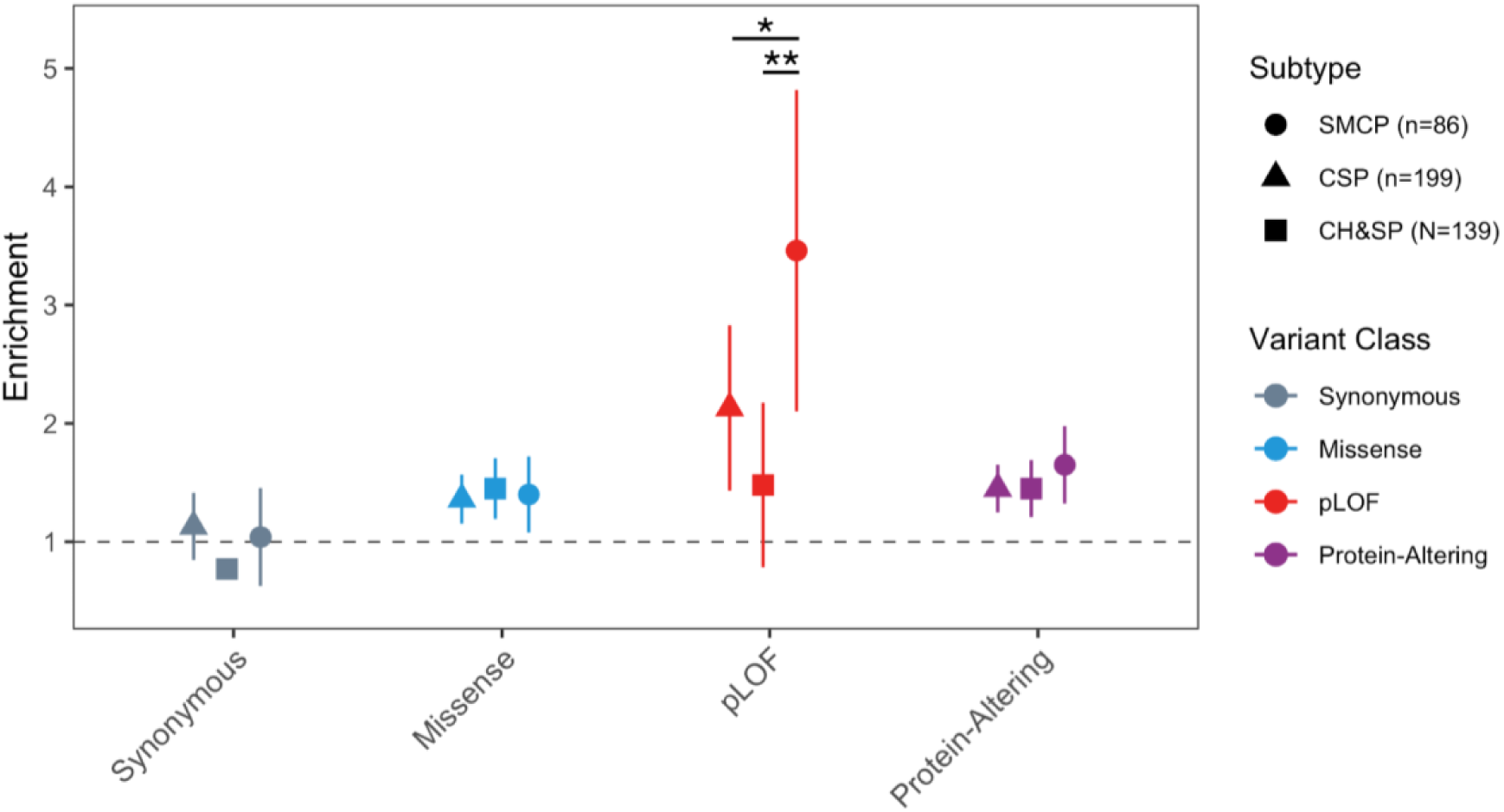
Exome-wide DN enrichment patterns across subtypes. There were no significant differences across any subtype for missense or protein-altering variants. SMCP probands were significantly more enriched compared to both CSP and CH&SP. The horizontal dotted line at 1 represents no enrichment (where observed = expected). *p=0.043, **p=0.003.

### Gene-specific analyses unveil known and previously unassociated candidate genes for CP

We first performed analysis on a per-gene basis to identify individual genes with a significant excess of DNs. In the full cohort, 4 genes reached exome-wide significance (p<1.3×10^-6^) for both PA and pLOF DNs: *SATB2* (PA p=1.24×10^-30^, pLOF p=1.64×10^-22^), *MEIS2* (PA p=2.15×10^-10^, pLOF p=4.97×10^-9^), *COL2A1* (PA p=4.91×10^-10^, pLOF p=2.84×10^-12^), and *ZC4H2* (PA p=2.30×10^-7^, pLOF p=1.15×10^-6^). Three additional genes were exome-wide significant for pLOF DNs only: *EFTUD2* (p=4.24×10^-8^), *KAT6B* (p=1.45×10^-7^), and *ANKRD11* (p=1.51×10^-7^) (**Figure 4**). Using a less conservative threshold correcting only for the number of genes with DNs (p<5.0×10^-5^), 7 additional genes were enriched for PA DNs including *IRF6 (*p=3.11×10^-6^), *MED13L (*p=7.48×10^-6^), *PRKCI (*p=8.57×10^-6^*), NEDD4L (*p=2.10×10^-5^), *EFTUD2 (*p=2.55×10^-5^), *ANKRD11* (p=3.65×10^-5^), and *STAG2 (*p=4.80×10^-5^) and 3 additional genes were enriched for pLOF DNs including *KMT2D* (p=1.98×10^-6^), *WDR26* (p=4.63×10^-5^), and *MYH3* (p=4.95×10^-5^). Of these 15 genes, all but *PRKCI* were previously associated with CP or OFCs.

**Figure 4:**
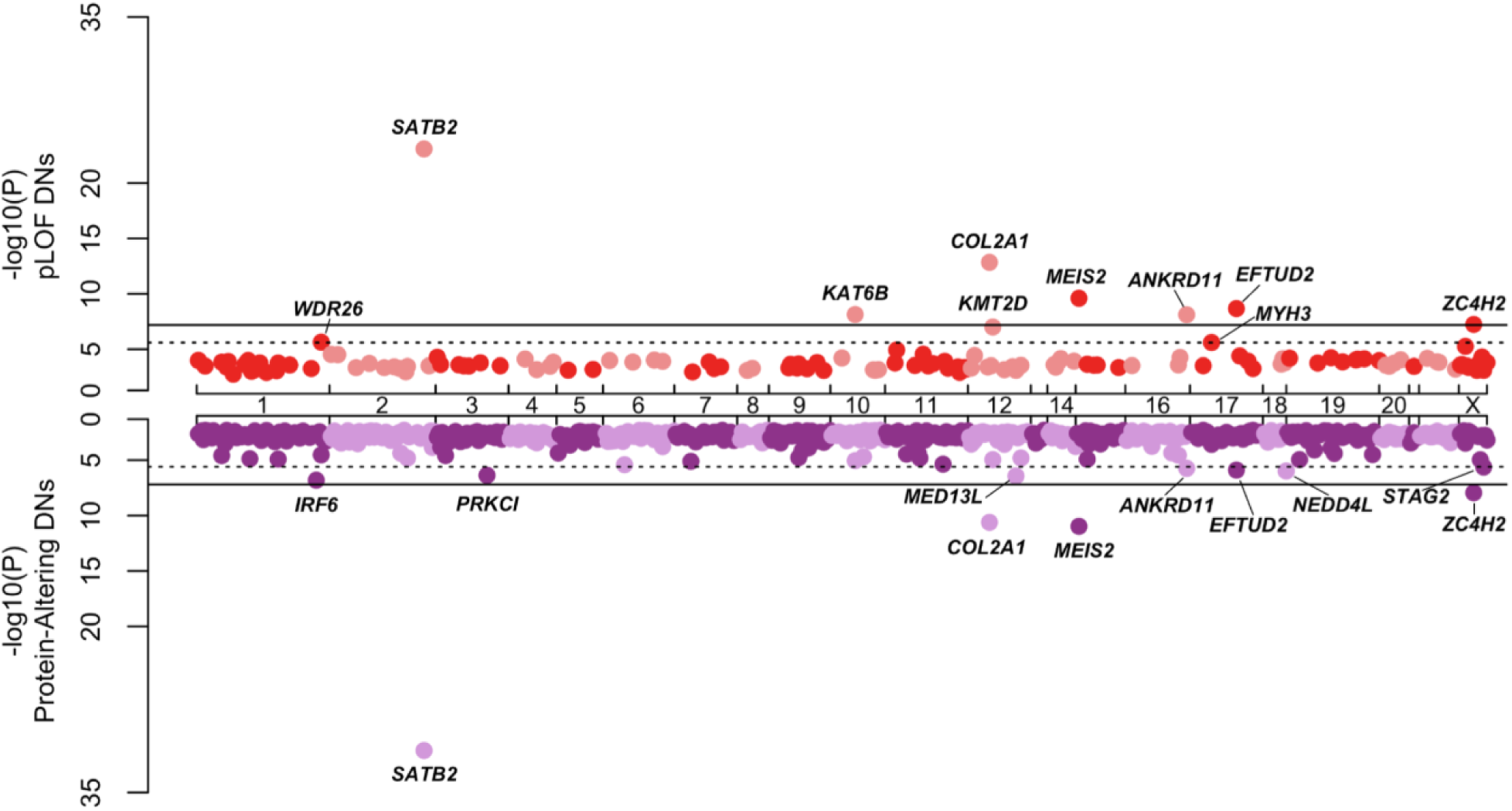
**Gene-specific analyses find exome-wide significance for putative loss-of-function (pLOF) (top) and protein-altering DNs (bottom).**

Enrichments in *ZC4H2*, *MED13L*, *ANKRD11*, and *KAT6B* were driven exclusively by DNs in syndromic probands whereas the *IRF6* enrichment was driven exclusively by nonsyndromic probands. The remaining genes had at least 1 DN identified in each group (**Table 1**). In total, there were 27 genes that shared PA DNs between both groups, though each was enriched to varying degrees due to differences in sample size (**Figure 5**, **Supplemental Table 6**). A total of 400 and 371 genes with DNs were found only in syndromic or isolated probands, respectively.

**Figure 5:**
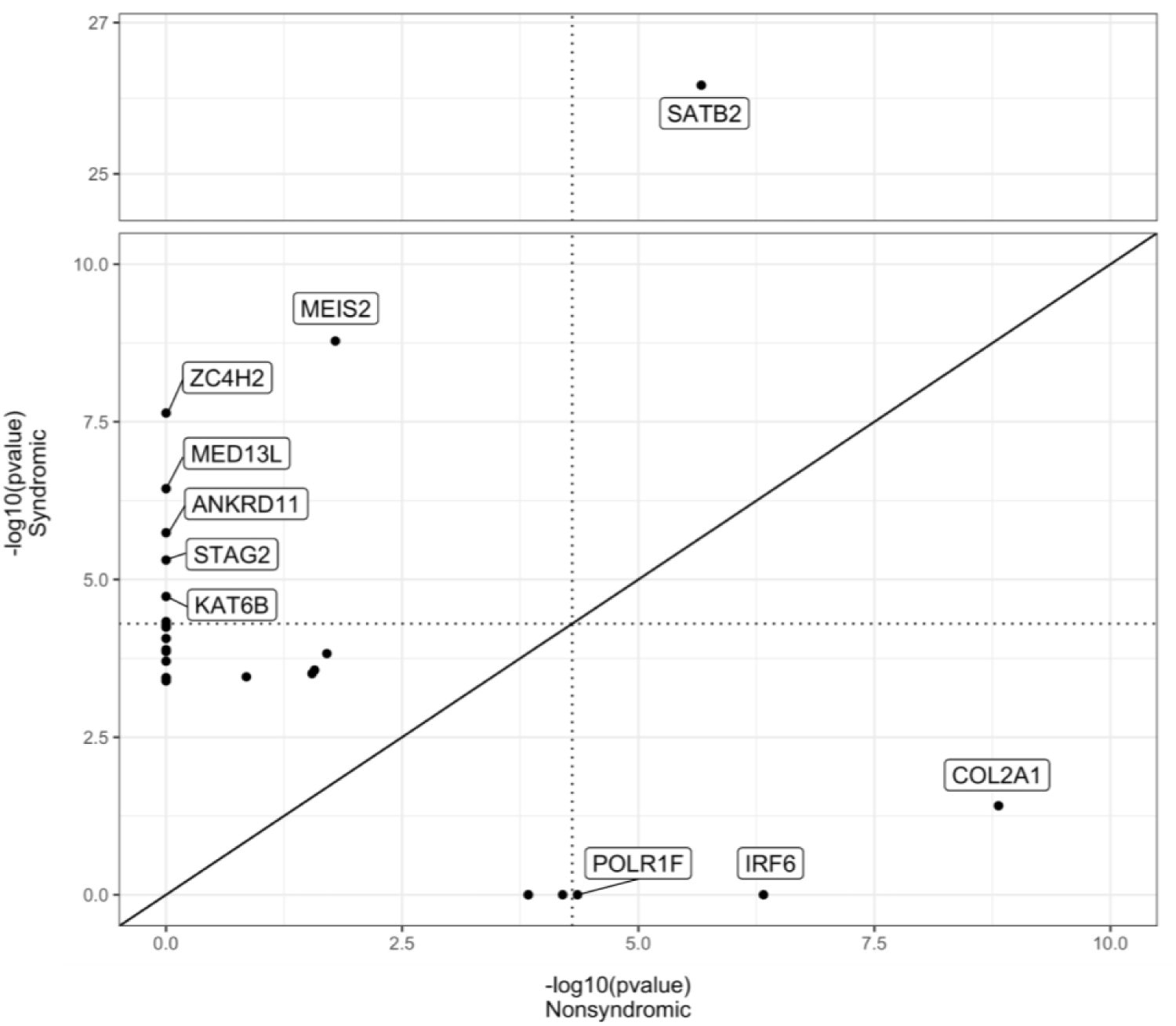
Comparison of DN enrichment by gene in syndromic versus nonsyndromic probands shows shared and distinct patterns. The dotted lines represent significant enrichment (p=5.0×10-5) and the solid line represents the expected value if enrichment were the same regardless of status.

**Table 1:** Top 15 genes by significance in combined CPSeq and DDD cohorts with individual contributions from syndromic and non-syndromic probands. Values in **bold** indicate exome-wide significance.

| Gene | All (N=816) |  | Syndromic (N=378) |  | Non-syndromic (N=434) |  |
| --- | --- | --- | --- | --- | --- | --- |
|  | Observed | p-value | Observed | p-value | Observed | p-value |
| *SATB2* | 14 | **1.24E-30** | 11 | **6.72E-27** | 3 | 2.15E-06 |
| *MEIS2* | 5 | **2.15E-10** | 4 | **1.65E-09** | 1 | 0.0161 |
| *COL2A1* | 6 | **4.91E-10** | 1 | 0.0387 | 5 | **1.53E-09** |
| *ZC4H2* | 3 | **2.30E-07** | 3 | **2.30E-08** | - | - |
| *IRF6* | 3 | 3.11E-06 | - | - | 3 | **4.73E-07** |
| *MED13L* | 4 | 7.48E-06 | 4 | **3.62E-07** | - | - |
| *PRKCI* | 3 | 8.57E-06 | 2 | 0.000149 | 1 | 0.0198 |
| *NEDD4L* | 3 | 2.10E-05 | 2 | 0.000273 | 1 | 0.0267 |
| *EFTUD2* | 3 | 2.55E-05 | 2 | 0.00031 | 1 | 0.0284 |
| *ANKRD11* | 4 | 3.65E-05 | 4 | 1.81E-06 | - | - |
| *KAT6B* | 3 | 0.000178 | 3 | 1.85E-05 | - | - |
| *KMT2D* | 4 | 0.000221 | 3 | 0.000349 | 1 | 0.141 |
| *WDR26* | 2 | 0.000595 | 2 | 0.000129 | - | - |
| *MYH3* | 2 | 0.00526 | - | - | 2 | 0.00154 |

We also assessed individual DNs in syndromic probands ascertained on CP (*i.e.,* syndromic probands in CPSeq) (**Supplemental Table 3**) and identified two variants that may explain each proband’s phenotype. First, we identified a variant in *CBL* (NM_005188:c.1228-2A>G) in an individual with CP, developmental delay, growth concerns, epilepsy, and an enlarged medial ventricle. In ClinVar (Accession: VCV000140455.3), this variant is classified as pathogenic/likely pathogenic for CBL syndrome (OMIM: 613563). Although epilepsy has not been reported for this syndrome, the remainder of the phenotypes are plausibly attributed to this finding. Similarly, we found a frameshift deletion in *RPL5* (NM_000969.5:c.46_47del) in a proband with CP, vascular ring anomaly, myopia, and ADHD. *RPL5* is associated with Diamond-Blackfan Anemia (OMIM: 612561), and though this specific variant has not been reported in ClinVar, it meets PVS1 (null variant), PS2 (confirmed *de novo*), and PM2 (absent in gnomAD v4.1.0) criteria therefore qualifying for pathogenic classification according to ACMG criteria. Though in-depth investigation of each DN is beyond the scope of this study, our findings highlight the value of searching beyond aggregate enrichment within a cohort, particularly where genotype supports the observed phenotypes.

Lastly, there were multiple genes with DNs found exclusively in specific CP subtypes (**Supplemental Table 7**). For example, 2 DNs each were found in *ARID1A* (p=1.89×10^-4^) and *TGFBR2* (p=1.50×10^-5^), all of which were found exclusively in probands with CHSP (n=139). Similarly, there were 3 *EFTUD2* (p=3.81×10^-7^) and 3 *PRKCI* (p=1.27×10^-7^) DNs all found in probands with CSP (n=199) specifically. Given the overall sample sizes, however, it is difficult to conclude whether our findings relate to the biological function of these genes or whether these patterns would change with larger samples.

### Genes associated with AD conditions featuring OFCs are significantly enriched for DNs

We next compared DN enrichment for a list of 418 genes with known associations with any OFC type (15) (**Supplemental Table 9**). Unsurprisingly, we observed higher and more significant enrichment within this restricted list of genes for PA DNs (5.89, p=1.45×10^-49^), including missense variants (3.3, p=3.80×10^-14^) and pLOF variants (24.3, p=3.23×10^-59^). When split by inheritance patterns, this list contained 178 genes associated with autosomal dominant (AD), 170 with autosomal recessive (AR), and 8 with X-linked conditions. The most pronounced findings were among genes related to AD conditions (10.2, p=1.01×10^-58^) as compared to AR conditions (1.84, p=0.017) (**Figure 6**), and there were no DNs in the limited number of X-linked genes. The difference in probands harboring DNs for AD versus AR OFCs was statistically significant for all PA DNs (p=3.83×10^-16^), with contributions from both missense (p=5.58×10^-6^) and pLOF DNs (p=4.57×10^-11^). This pattern remained true regardless of syndromic status or subtype classification, though we did not statistically test more stratified comparisons (**Supplemental Table 10**, **Supplemental Figures 2 & 3**).

**Figure 6:**
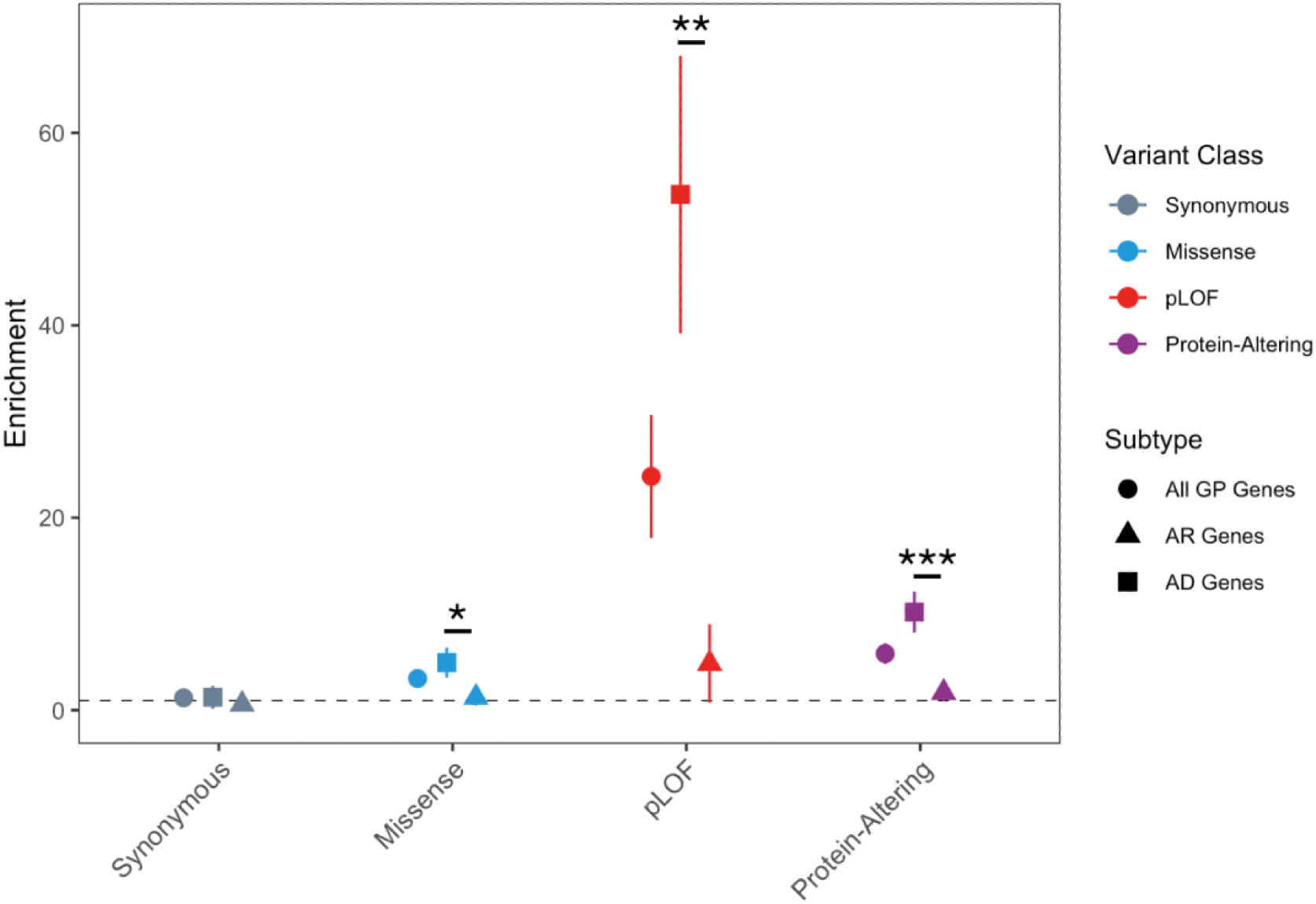
CP probands are strongly enriched for OFC-associated genes implicated in autosomal dominant conditions. Enrichment is shown for all genes in the panel and stratified by the mode of inheritance when known for disease-associated genes. The horizontal dotted line at 1 represents no enrichment (observed = expected value). Variants classes are represented by the following colors: gray=synonymous, blue=missense, red=pLOF, purple=protein-altering. *p=5.58×10^-6^, **p=4.57×10^-11^, ***p=3.83×10^-16^.

In aggregate, 11.4% of all DNs (126/1102) and 13.3% (116/875) of PA DNs belonged to genes in this list. When stratified by inheritance patterns, the distribution of these PA DNs was consistent with our enrichment findings: 79.3% (92/116) were associated with AD, 13.8% (16/116) with AR, and 6.0% (7/116) with unspecified conditions. Further, we found that 6.3% (52/816) of probands harbored a pLOF DN in a gene associated with AD OFCs, and 16 of these were found in nonsyndromic individuals. From a clinical standpoint, these findings support genetic testing for any individual with CP, regardless of syndromic status or family history.

### Gene Ontology Suggests Differences in Underlying Mechanisms of Syndromic and Nonsyndromic CP

In order to understand the biological implications of DNs in CP and its phenotypic groupings, we compared gene ontology results (all with adjusted p-value <0.05) for biological processes (BP), cellular component (CC), and molecular function (MF) categories using gProfiler (25). As a positive control, we first compared enrichment of synonymous DNs in syndromic or nonsyndromic probands. There were no enriched terms for the nonsyndromic group, and only two terms (MF: protein-binding and CC: endomembrane system) for syndromic probands, indicating very little specificity as would be expected. We then compared enrichment for PA DNs between groups, finding notable differences in the so-called “driver terms” as highlighted by gProfiler, as well as the combined results of all enriched terms (**Supplemental Table 8**). For genes with PA DNs in nonsyndromic probands, the driver terms included broad biological functions including “anatomical structure morphogenesis”, “biological regulation”, “organelle organization”, “actin filament-based process”, and “response to stimuli”, molecular functions including “ion binding”, “ATP hydrolysis activity”, and “protein binding”, and a range of cellular components that also included several cytoskeletal terms. In contrast, genes from the syndromic probands included MF terms such as “histone modifying activity”, “DNA binding”, and “transcriptional co-regular activity”. There was also no overlap in BP driver terms for syndromic probands with results including “chromatin remodeling”, “DNA metabolic process”, “cell surface receptor signaling pathway”, and “double strand break repair”. Similarly, CC terms did not generally overlap between syndromic and nonsyndromic gene lists and, for syndromic-associated genes, included “cell projection”, “cytosol”, “cell junction”, “centriole”, “somatodendritic compartment”, and “U2AF complex”. In general, these findings are consistent which would be expected from a dataset of syndromic probands featuring CP and NDDs; however, they also highlight the potential mechanistic differences in development of CP with or without additional phenotypes. As has been reported previously (16), we observed enrichment in terms related to chromatin remodeling in our syndromic group, along with DNA binding and transcriptional activity. Further investigation into genes belonging to these pathways is warranted.

### Distinct cell types are enriched for syndromic versus nonsyndromic CP probands

Single nucleus RNAseq data allows identification of specific sets of genes, or marker genes, that are highly expressed within cell populations. We next wanted to know if any specific cell types were overrepresented in our DN dataset as this may allow a more granular understanding of the key cells at play in CP development. For simplicity, we only report enrichment for the PA group in the following section, though all DN class enrichment is available in the supplemental data.

We first looked at marker genes across 10 clusters derived from the secondary palate of mice at E15.5 (the time of palatal shelf fusion) with an FDR of <0.01 (**Supplemental Table 11**) (26). From the 2,647 total marker genes, there were 189 unique genes (containing 237 total DNs). In the full cohort, there were 6 clusters significantly enriched (p<0.005, correction for 10 clusters): chondrocyte progenitor cells (3.62, p=2.94×10^-5^), early osteocyte progenitor cells (3.72, p=3.11×10^-7^), late osteocyte progenitor cells (1.9, p=2.75×10^-3^), endothelium (1.88, p=2.24×10^-5^), epithelium (1.64, p=1.17×10^-3^), and mesenchyme (2.0, p=2.82×10^-3^) (**Table 2**, **Figure 7A**, **Supplemental Table 12**). When stratified by syndromic status, we found chondrocyte progenitor cells and endothelium remained significantly enriched solely in our nonsyndromic group, whereas early osteocyte progenitor cells, late osteocyte progenitor cells, and mesenchyme remained significant only in the syndromic group (**Figure 7A**, **Supplemental Figure 4A**). The epithelium was not significantly enriched in either group alone.

**Figure 7:**
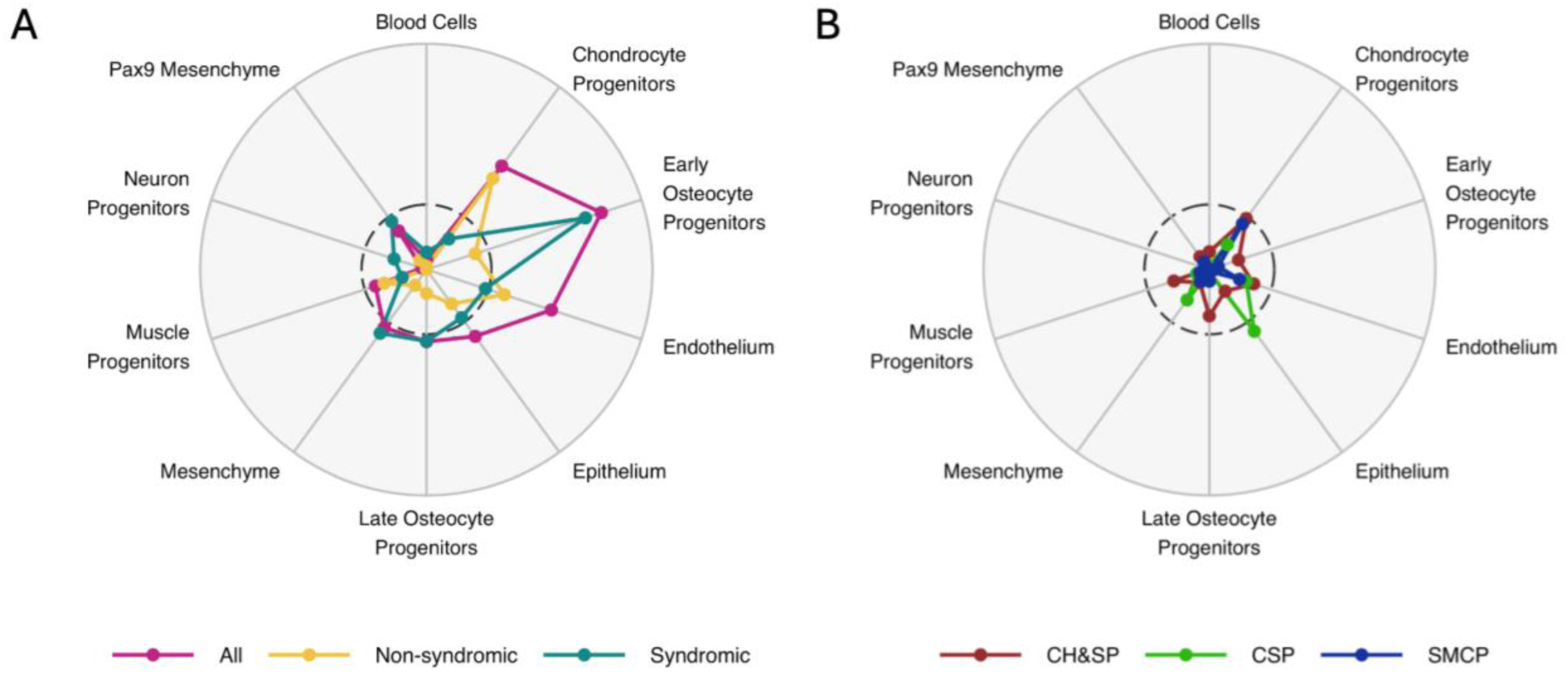
Sets of marker genes from mouse palate at E15.5 are differentially enriched based on syndromic status and CP subtype. Radar plots for −log10(pvalue) showing significant enrichment by cluster by A) syndromic status and B) CP subtype. The center dotted line represents p<0.005 and the outermost line represented p<1×10^-8^.

**Table 2:**
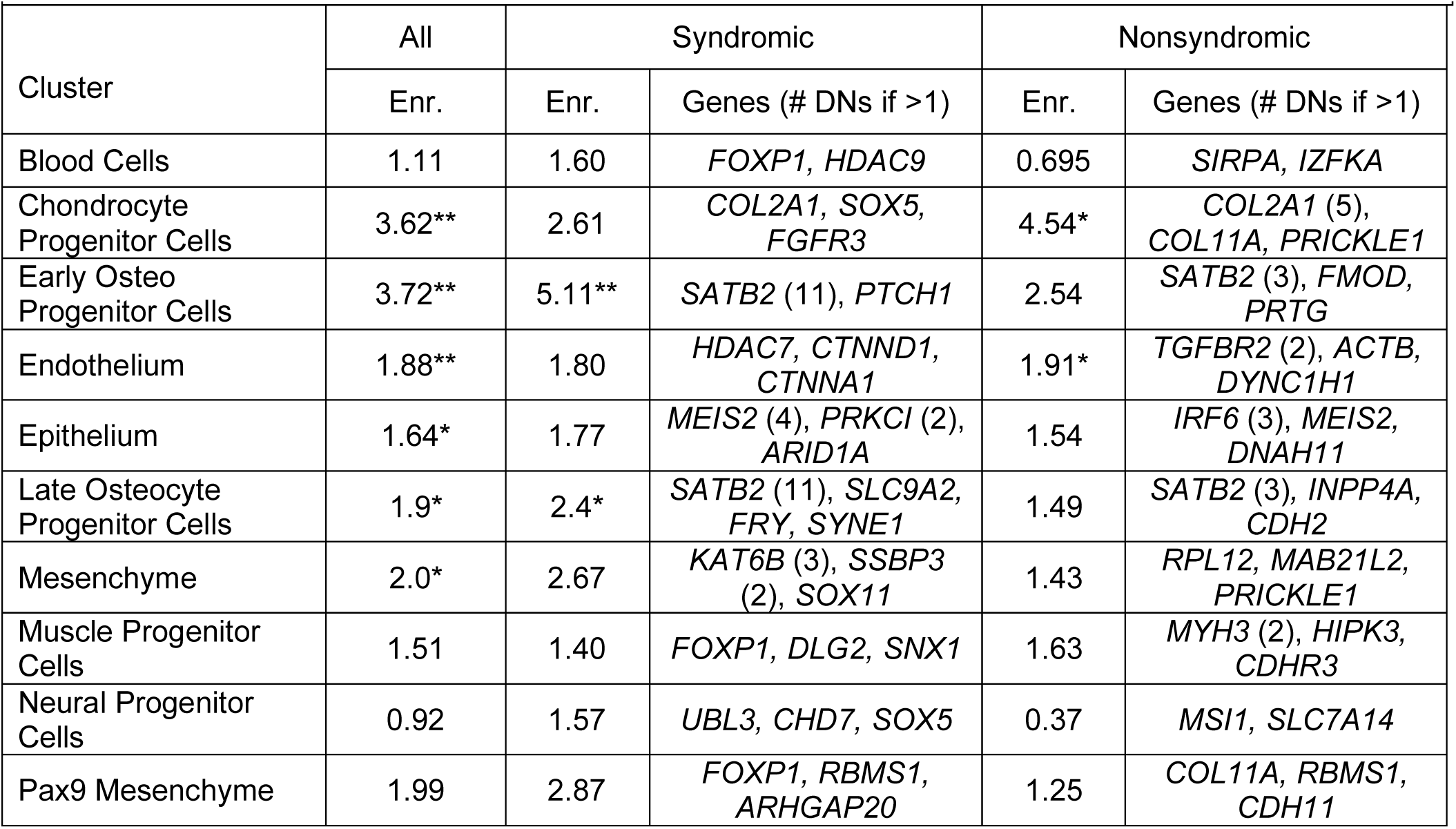
Enrichment and select genes for protein-altering DNs in 10 clusters from the mouse secondary palate at embryonic day 15.5. Enr=enrichment. *Enrichment is significant with p<5×10^-3^ **Enrichment is significant with p<5×10^-5^

When looking at the specific genes within these clusters, we found varied patterns of DN burden. There were 5 *COL2A1* DNs within the nonsyndromic probands, accounting for 50% of the observed variants in the chondrocyte progenitor cluster. In contrast, among the 28 DNs found within genes of the endothelium, only two (*TGBGR2* and *DYNC1H1*) harbored multiple DNs, with 2 each. Similarly, in the syndromic group, *SATB2* variants accounted for 79% (11/14) of DNs in both early and late osteocyte progenitors, whereas in the mesenchyme, only two genes had more than one DN out of 13 total (*KAT6B*, n=3; *SSBP3*, n=2). Collectively, these results suggest while some genes are individually enriched for DNs, such as *COL2A1* or *SATB2*, disruption of broader processes (rather than a specific gene) are also contributing to CP pathophysiology. For example, 24% (6/25) of the endothelium marker genes with DNs in nonsyndromic probands are part of the actin cytoskeleton; thus, deeper investigation into more general pathways and/or cell components may lead to more discoveries.

As with other evaluations, we also compared cluster enrichment by CP subtype to identify specific cell types possibly reflecting phenotypic heterogeneity. However, the only cluster with any significant enrichment was the epithelium in CSP (2.34, p=1.95×10^-3^) (**Figure 7B**, **Supplemental Table 12, Supplemental Figure 4B**). There were 16 DNs in these cluster marker genes: 3 were within *PRKCI* and the remainder had only 1 DN per gene. Although we did not identify any significant patterns by subtype, future studies with larger sample sizes may provide more insightful and stronger evidence for the trends we observed.

We also considered that investigation into earlier cell origins could reveal differences between CP subgroups, as the mouse data was derived from palatal tissue specifically at the time of osteogenesis. As such, we next evaluated DNs in marker genes from a publicly available scRNAseq dataset derived from human embryos at post-conceptional weeks 3-5 and clustered into 14 main cell lineages (27). In this dataset, there were 952 total marker genes (**Supplemental Table 13**), of which 51 unique genes contained 65 total DNs. Four clusters were significantly enriched (p<3.57×10^-3^, correction for 14 clusters), including epithelium (4.91, p=1.01×10^-5^), intermediate mesoderm (4.31, p=1.71×10^-5^), dermomyotome (5.63, p=1.74×10^-5^), and sclerotome (3.82, p=3.96×10^-5^), though the lateral plate mesoderm was barely shy of our cutoff (3.03, p=3.59×10^-3^) (**Table 3, Supplemental Table 14**, **Supplemental Figure 4**). With this dataset, genes for the epithelium, dermomyotome, and sclerotome were enriched for DNs in nonsyndromic probands whereas the intermediate mesoderm and lateral plate mesoderm were enriched for DNs in the syndromic probands (**Figure 8A**, **Supplemental Figure 5A**). However, as with the mouse data, *COL2A1* was a key driver in all enriched clusters for nonsyndromic probands, making up 56% (5/9), 71% (5/7), and 71% (5/7) of the DNs in epithelium, dermomyotome, and sclerotome, respectively. A similar pattern for syndromic probands was found, with *MEIS2* accounting for 44% (4/9) intermediate mesoderm and 67% (4/6) lateral plate mesoderm DNs. Unlike the mouse data, there were no enriched clusters lacking major driver genes.

**Figure 8:**
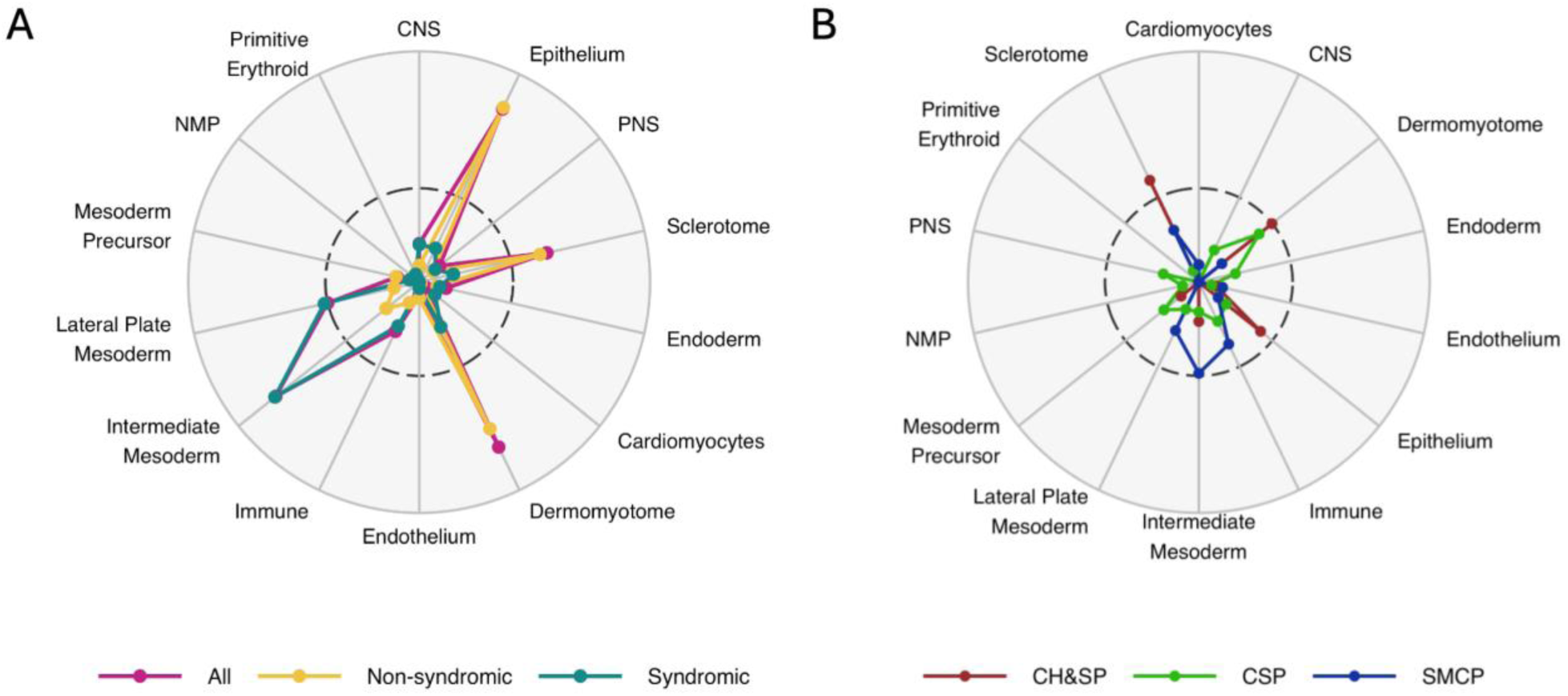
Sets of marker genes from human embryos are differentially enriched based on syndromic status and CP subtype. Radar plots for −log10(pvalue) showing significant enrichment by cluster by A) syndromic status and B) CP subtype. The center dotted line represents p<0.005 and the outermost line represented p<1×10^-6^.

**Table 3:** Enrichment and select genes for protein-altering DNs in 14 clusters from human embryos at post-conceptional weeks 3-5. *Enrichment is significant with p<5×10^-3^ **Enrichment is significant with p<5×10^-5^

| Cluster (by lineage) | All | Syndromic |  | Nonsyndromic |  |
| --- | --- | --- | --- | --- | --- |
|  | Enr. | Enr. | Genes | Enr. | Genes |
| Cardiomyocyte | 0.964 | 1.56 | *TTN* | 0.453 | *TTN* |
| CNS lineage | 2.05 | 2.65 | *ELAVL1, PBX1, SOX11* | 1.54 | *CDH2, KIF21A* |
| Dermomyotome | 5.63** | 3.64 | *COL2A1, RPL2, RPS5* | 7.41* | *COL2A1 (5), RPL12, RPS6* |
| Endoderm | 1.77 | 1.91 | *CDKN1C, LAMB1* | 1.67 | *SERPINA1, DSC2* |
| Endothelium | 1.21 | 0.868 | *SLC9A3R2* | 1.51 | *ANKRD37, ARPC1B* |
| Epithelium | 4.91 | 2.65 | *COL2A1, RPL2, RPS5* | 6.92** | *COL2A1 (5), RPL12, PRTG* |
| Immune | 2.75 | 3.56 | *CTSZ, FCGRT, MAF* | 2.07 | *APRBC1 ACTB* |
| Intermediate mesoderm | 4.31** | 6.44** | *MEIS2 (4), CDKN1C, PAX8* | 2.49 | *MEIS2, RBMS1, PEG3* |
| Lateral plate mesoderm | 3.03* | 4.36* | *MEIS2 (4), CDKN1C, FOXP1* | 1.9 | *MEIS2, DSC2, PEG3* |
| Mesoderm precursor | 1.71 | 1.23 | *FBL* | 2.15 | *RPS6, SMC6* |
| Neuromesodermal | 0.535 | - | - | 1.01 | *RPS6* |
| PNS lineage | 1.7 | 1.84 | *RPS2, BCAR3* | 1.6 | *YBX1, ARPC1B* |
| Primitive erythroid | 0.505 | 1.09 | *HK1* | - | - |
| Sclerotome | 3.82* | 2.47 | *CCD32, COL2A1, FBN2* | 5.03* | *COL2A1 (5), CDH11, SNAI1* |

We found a single enriched cluster when stratified by CP subtype (**Figure 8B**, **Supplemental Figure 5B**), with the sclerotome remaining significant for CH&SP probands, with 2 DNs in in *COL2A1*, and 1 each for *CDH1* and *SNAI1*. We noted fewer overall DNs overlapping with the human data (5.3% human versus 7.1% mouse), which may suggest more specificity for DNs in genes in the palate leading to CP but may also be due to factors not related to biology such as statical analysis methods or other technical differences.

In summary, both datasets were enriched for DNs in this cohort, with distinct patterns emerging primarily between syndromic and nonsyndromic CP probands. Although larger samples sizes are needed for further validation, investigation into variants within other genes of the enriched cell clusters may provide additional insight into CP etiology.

## Discussion

Here we found exome-wide enrichment in a large-scale investigation of coding DNs in 816 CP trios representing multiple CP subtypes and a broad phenotypic spectrum. There was a significantly higher burden of DNs in syndromic probands, which was not unexpected given that *de novo* variation is not under selective pressure and therefore can result in more severe phenotypes. Interestingly, although there was no significant difference in the DN rate between males and females within our cohort in any group, there was deviation in the M:F ratio in probands classified as syndromic when compared to reported ratios from EUROCAT. We suspect this observation is a result of ascertainment. The majority of our syndromic probands were ascertained on NDDs as part of the DDD study, which disproportionally affect males (29, 30).

A second possible effect of ascertainment differences between DDD and CPSeq was the lack of DNs in genes frequently associated with syndromic forms of CP that are not characterized by intellectual disability or neurological phenotypes. Of note, there were no syndromic probands harboring DNs in *IRF6* (31), *GRHL3* (32), or *MYH3* (33)—these were only found in our presumed nonsyndromic individuals. These results likely indicate differing mechanisms underlying syndromic CP featuring NDDs versus other congenital anomalies, illustrating care must be taken to not extrapolate the “syndromic” results presented here to any and all syndromic forms of CP. The presence of PA DNs in the nonsyndromic cohort in genes typically associated with syndromic OFCs raises other important issues facing human genetics including understanding variable expressivity versus the limitations of phenotyping. Some of these “syndromic” genes with DNs in the nonsyndromic cohort included *COL21A* (34), *SATB2* (35), and *MEIS2* (36). While variable expressivity has been documented for *IRF6* (31, 37), *GRHL3* (9, 32), and *COL2A1* (38, 39), this is less true for the other genes mentioned here. It is important to note, however, that our nonsyndromic probands are presumed so—some were recruited in infancy during which additional features, particularly NDDs, would not have yet been apparent. Still, our findings could suggest an expansion of the phenotypic spectrum for *SATB2*, *MYH3,* and *MEIS2* to include isolated CP.

Previous work by Wilson et al. found pathogenic/likely pathogenic variants in syndromic CP probands were overrepresented in genes involved in chromatin remodeling, which has historically been enriched in NDDs. This is similar to our findings, where syndromic probands were enriched for both chromatin remodeling and histone modifying activity, as well as DNA binding, transcriptional co-regular activity, and double stranded break repair. It is important to note, however, there is much overlap in the data between studies, as many of their samples also came from DDD. Broadly, however, the overlaps and distinctions between gene ontology for syndromic and nonsyndromic probands can provide clues to etiologic differences. For example, nonsyndromic probands were enriched in processes related to actin filament binding, which are key in epithelial cell dynamics (40, 41). Perhaps failure of cell migration and/or adhesion of epithelial cells during palatal development is more likely to result in a failure of palatal shelf fusion than additional systemic anomalies. This idea is further supported by the human snRNAseq enrichment, in which nonsyndromic probands were enriched for DNs in marker genes from the epithelium cluster. Conversely, it would then make sense that disruption of processes affecting broader cell types, such as histone modification or transcriptional activity, could then result in broader effects as observed in syndromic probands.

Understanding the genetic underpinnings of CP subtypes remains a challenge. It could be hypothesized that CSP is a less severe version of CH&SP. The current data do not support this idea, as there were no differences in the rate of DNs in any class of variant between the two. In fact, although not significantly different, there was a higher enrichment of pLOF DNs for CSP than for CH&SP, suggesting a heterogenous genetic architecture underlying similar phenotypic heterogeny. We also aimed to identify differences in specific cell types or biological processes and found two enriched clusters by subtype: CH&SP in the sclerotome from the human data and CSP in the mouse epithelium cluster. Although the sclerotome does not directly contribute to craniofacial development, it does similarly give rise bony structures (the vertebral column) and associated soft tissues (intervertebral discs, meninges) (42). Therefore, it would be unsurprising to observe an enrichment for CH&SP PA DNs if these genes play similar roles in palatal development, but further study is needed to substantiate such speculation. We also found probands with CSP were enriched for DNs in genes from the mouse epithelium cluster. This may indicate a higher likelihood for CSP with disruption of genes in the epithelium, but we also know that genes highly expressed in this tissue, such as *IRF6* and *MEIS2*, are not specifically associated with any CP subtype, so these associations must be interpreted cautiously.

Taken altogether, these data imply that there is not currently a single list of genes that represents all individuals with CP, and care should be taken when creating lists of “cleft palate genes.” We highlight the utility of combined analysis of all classifications of CP to identify new candidate genes for CP and to expand the phenotypic spectrum for others. In aggregate, we show there are differences between syndromic or nonsyndromic probands, though on an individual level we find these distinctions are less clear cut. When considering this in a larger clinical picture, genetic testing for any individual born with a CP regardless of the presence of additional clinical features may be fruitful—in fact, we found that 6.3% (52/816) of probands had a pLOF DN in an AD OFC-associated gene, 16 of which were in presumed nonsyndromic individuals. Although more detailed curation of variant pathogenicity is needed for the full list of DNs, this preliminary finding suggests that the clinical diagnostic yield for CP in this cohort may be similar to recent reports (15). Future studies focused on the spectrum of phenotypes in genes associated with OFC syndromes, deeper exploration of subtype genetic risks, and the contribution of rare inherited variants are warranted to better understand palatogenesis and the genetic architecture of CP.

## Declaration of interests

The authors declare no competing interests.

## Data Availability

Sequence and phenotype data is available from the Database of Genotypes and Phenotypes (dbGaP, ncbi.nlm.nih.gov) under study accession # phs002220.v1.p1 (CPSeq probands) and accession # phs001168.v2.p2 (US, European proband). Additional information for probands in the GMKF cohort can be viewed at https://kidsfirstdrc.org/studies/.

## Acknowledgements

We are very thankful for participants, their families, and colleagues who have made this research possible. Sequencing services for CPSeq were provided by the Center for Inherited Disease Research (CIDR). CIDR is fully funded through a federal contract from the National Institutes of Health to The Johns Hopkins University, contract number HHSN268201700006I.

Additional whole genome sequencing was funded by National Institutes of Health (NIH) grants: X01-HG010835 (EJL, MLM), X01-HL0132363 (MLM), and X01-HD100701 (EJL, JCM, MLM).

Patient recruitment, assembly of phenotypic information, and data analysis were supported by National Institutes of Health (NIH) grants: F31-DE032588 (KR), R01 DE027983 (EJL), R01-DE028342 (EJL), R01-DE030342 (EJL), R01-DE028300 (AB), R00-DE024571 (CJB), U54GM133807 (CJB), U54GM133807 (CJB), R01-DE008559 (JCM), R01-DE016148 (MLM, SMW), R01-DE008559 (JCM, MLM), R01-DE032122 (MLM), R01-DE0332319 (MLM, EJL, SMW), R01-DE011931 (JTH), and R01-DE031261 (HB). We thank the California Department of Public Health, Maternal Child and Adolescent Division, for providing data for these analyses. This work was supported by the Centers for Disease Control and Prevention, Centers of Excellence No. U01-DD001033 (GMS).

## Author contributions

This study conceptualized by EJL and KRR. Resources were contributed by THB, AB, CJB, KFD, JTH, LMU, JCM, JEP, GMS, SMW, ECL, HB, MLM, and EJL. Data curation was performed by KRR, SC, JEP, DJC, KFD, and HB. Data analysis, investigation, and visualization were performed by KRR under supervision of DJC, MPE, and EJL. The originally manuscript was drafted by KRR. All authors contributed to critical review and approval of the final draft.

## Notes

### Competing Interest Statement

The authors have declared no competing interest.

### Author Declarations

The IRB of Emory University gave ethical approval for this work.

