## Supplemental_Figures for "Distinguishing syndromic and nonsyndromic cleft palate through analysis of protein-altering de novo variants in 816 trios"

Supplemental Figure 1: DN distribution A) per trio and B) by variant class.

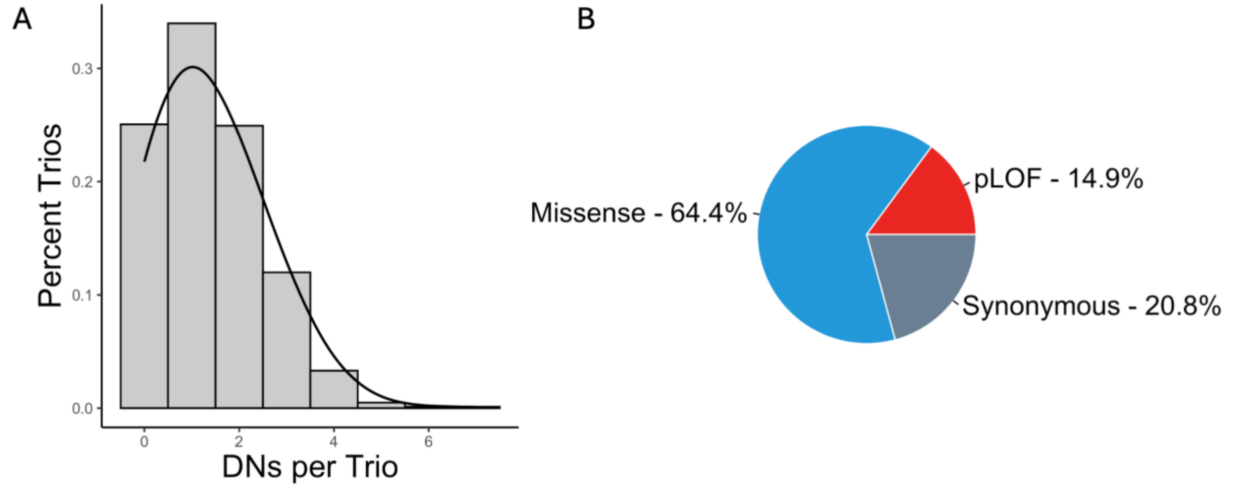


Supplemental Figure 2: DN enrichment for OFC associated genes by syndromic status. The horizontal dotted line at 1 represents no enrichment (where observed = expected).

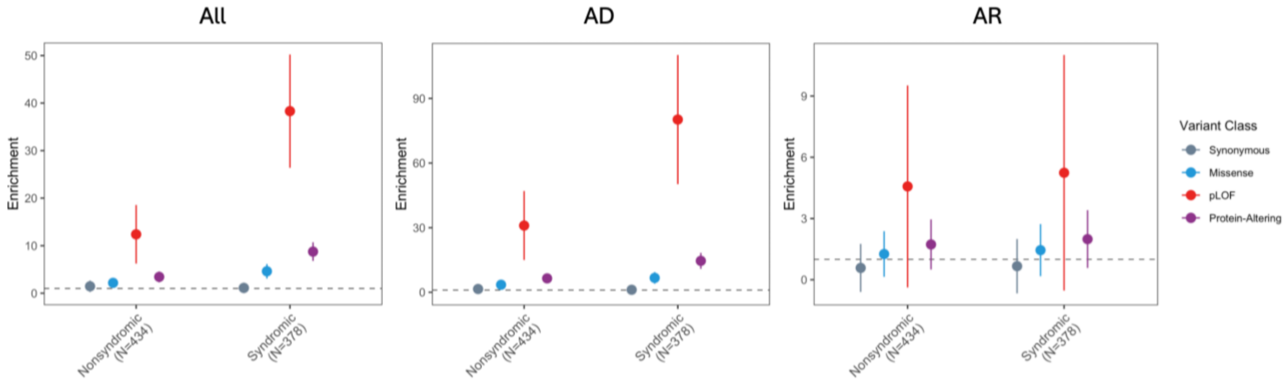


Supplemental Figure 3: DN enrichment for OFC associated genes by CP subtype. The horizontal dotted line at 1 represents no enrichment (where observed = expected).

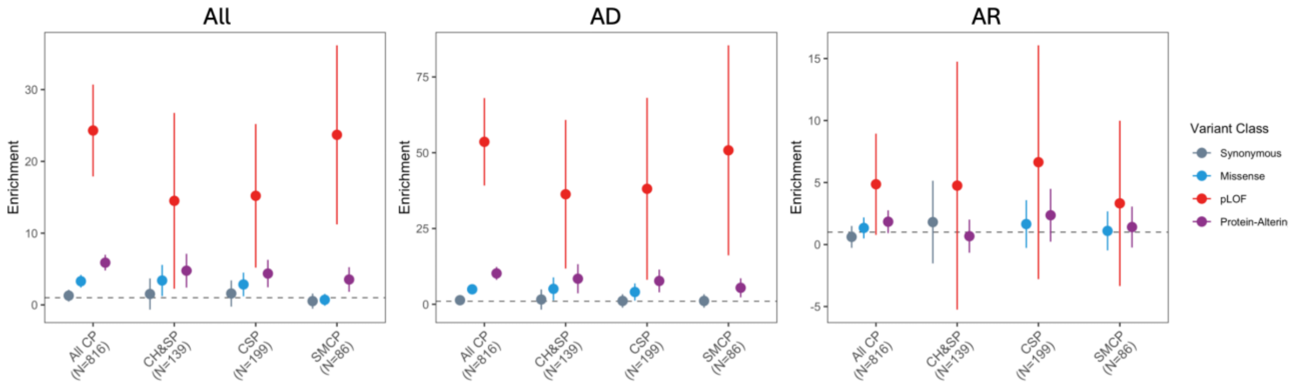

Supplemental Figure 4: Enrichment in gene sets from snRNAseq clusters in the mouse palate at E15.5 by variant class for A) syndromic status and B) CP subtype.

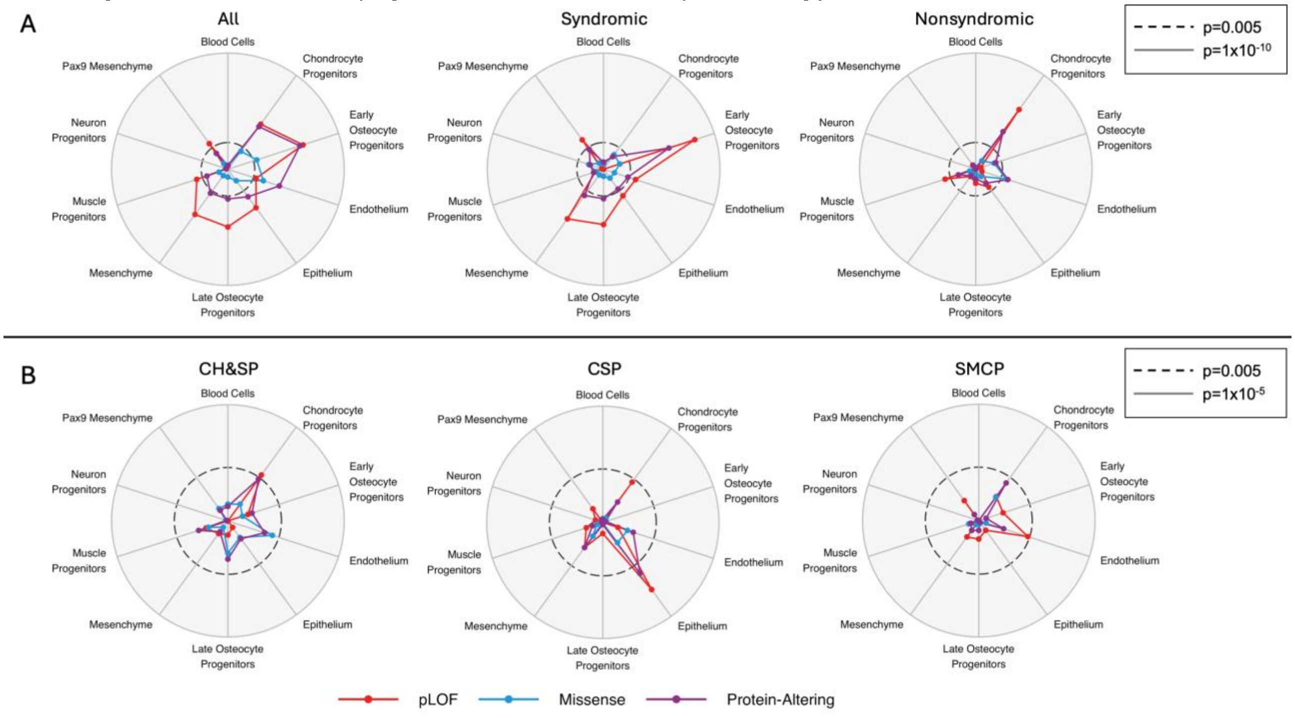


Supplemental Figure 5: Enrichment in gene sets from snRNAseq clusters in human embryos by variant class for A) syndromic status and B) CP subtype.

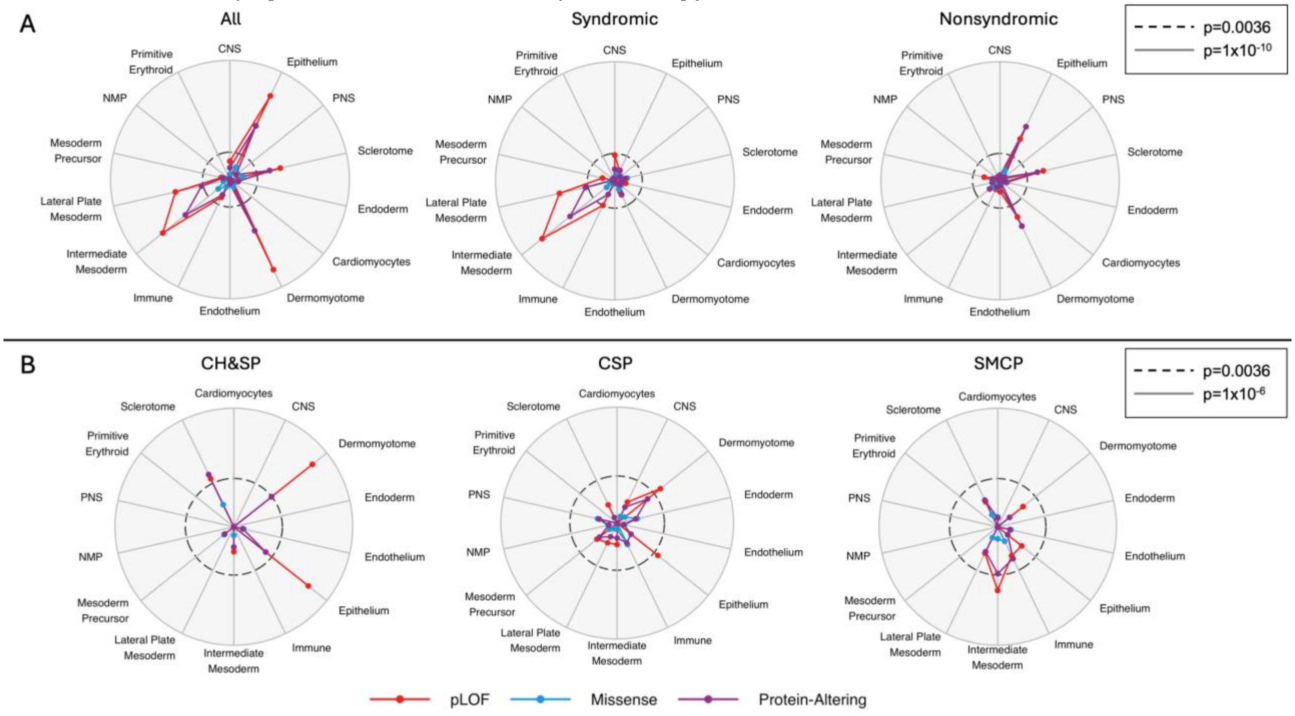
